# Improved Detection of Air Trapping on Expiratory Computed Tomography Using Deep Learning

**DOI:** 10.1101/2020.11.17.20229344

**Authors:** Sundaresh Ram, Benjamin A. Hoff, Alexander J. Bell, Stefanie Galban, Aleksa B. Fortuna, Oliver Weinheimer, Mark O. Wielpütz, Terry E. Robinson, Beverley Newman, Dharshan Vummidi, Aamer Chughtai, Ella A. Kazerooni, Timothy D. Johnson, MeiLan K. Han, Charles R. Hatt, Craig J. Galban

**Affiliations:** Department of Radiology, University of Michigan, Ann Arbor, MI 48109, USA; Department of Biomedical Engineering, University of Michigan, Ann Arbor, MI 48109, USA; Department of Diagnostic and Interventional Radiology, University Hospital of Heidelberg, 69120 Heidelberg, Germany; Translational Lung Research Center, Heidelberg (TLRC), German Lung Research Center (DZL), 69120 Heidelberg, Germany; Center of Excellence in Pulmonary Biology, Department of Pediatrics, Stanford University School of Medicine, Stanford, CA 94304, USA; Department of Pediatric Radiology, Lucile Packard Children’s Hospital at Stanford, Stanford, CA, 94305, USA; Department of Internal Medicine, Michigan Medicine, University of Michigan, Ann Arbor, MI 48109, USA; Department of Biostatistics, University of Michigan, School of Public Health, Ann Arbor, MI 48109, USA; Imbio LLC, Minneapolis, MN 55413, USA

**Author notes:** Corresponding author: Craig J. Galbán, Ph.D., Director of Preclinical Imaging & Computational Analysis, UM Rogel Cancer Center, Professor, Department of Radiology, Adjunct Associate Professor, Department of Biomedical Engineering, University of Michigan, 109 Zina Pitcher Place, Ann Arbor, MI 48109-2200; Phone (google).

**Keywords:** Machine learning, air trapping, cystic fibrosis, computed tomography, convolutional neural networks, artificial intelligence

## Abstract

**Background:** Radiologic evidence of air trapping (AT) on expiratory computed tomography (CT) scans is associated with early pulmonary dysfunction in patients with cystic fibrosis (CF). However, standard techniques for quantitative assessment of AT are highly variable, resulting in limited efficacy for monitoring disease progression.

**Objective:** To investigate the effectiveness of a convolutional neural network (CNN) model for quantifying and monitoring AT, and to compare it with other quantitative AT measures obtained from threshold-based techniques.

**Materials and Methods:** Paired volumetric whole lung inspiratory and expiratory CT scans were obtained at four time points (0, 3, 12 and 24 months) on 36 subjects with mild CF lung disease. A densely connected CNN (DN) was trained using AT segmentation maps generated from a personalized threshold-based method (PTM). Quantitative AT (QAT) values, presented as the relative volume of AT over the lungs, from the DN approach were compared to QAT values from the PTM method. Radiographic assessment, spirometric measures, and clinical scores were correlated to the DN QAT values using a linear mixed effects model.

**Results:** QAT values from the DN were found to increase from 8.65% ± 1.38% to 21.38% ± 1.82%, respectively, over a two-year period. Comparison of CNN model results to intensity-based measures demonstrated a systematic drop in the Dice coefficient over time (decreased from 0.86 ± 0.03 to 0.45 ± 0.04). The trends observed in DN QAT values were consistent with clinical scores for AT, bronchiectasis, and mucus plugging. In addition, the DN approach was found to be less susceptible to variations in expiratory deflation levels than the threshold-based approach.

**Conclusion:** The CNN model effectively delineated AT on expiratory CT scans, which provides an automated and objective approach for assessing and monitoring AT in CF patients.

## INTRODUCTION

High resolution CT is an integral tool for the treatment and management of patients with diffuse lung disease [1]. High resolution CT lung imaging provides high tissue-air contrast and resolution facilitating disease detection and characterization, and assessment of disease progression across a variety of obstructive and restrictive lung diseases. Mosaic attenuation on CT images is defined as a CT pattern that “appears as patchwork of regions of differing attenuation that may represent (i) patchy interstitial disease, (ii) obliterative small airways disease, or (iii) occlusive vascular disease” [2]. In the context of small airways disease, mosaic attenuation represents air trapping (AT) “secondary to bronchial or bronchiolar obstruction” that produces focal zones of decreased attenuation on expiratory CT imaging [2].

A mosaic attenuation pattern on expiratory CT scans due to AT is a common feature in many pulmonary conditions with airway obstruction [3]. Computational techniques that are fully automated have been developed to quantify the extent of AT on expiratory CT, which may improve the detection of AT across a diverse range of radiologists in practice. The most extensively used method is quantification of low attenuation areas using a Hounsfield unit (HU) threshold-based approach, which defines areas at or below a static attenuation value as AT. This approach was first applied to emphysema, and has been pathologically validated [4-7]. This same strategy has been used to quantify AT in small airway disease associated with chronic obstructive pulmonary disease (COPD), asthma, and obliterative bronchiolitis [8-13]. In many cases radiographically identified AT is not captured by threshold-based techniques. To address this limitation in automated AT quantification, adaptive techniques that calculate personalized thresholds [9] have been developed. Although simple to use and readily available, these attenuation threshold-based methods are known to be sensitive to scanner noise, reconstruction kernel, and the level of expiration at which the CT images were obtained [14]. This can create a discordance between the algorithm output and visual findings of AT by radiologists on the same chest CT images.

Recently, the availability of large amounts of data and significant computational power have rapidly increased the popularity of machine learning (esp. deep learning) approaches [15]. Convolutional neural networks (CNNs) [16] have been investigated in many image analysis tasks [17-19] and radiological applications [20, 21], and have outperformed the state-of-the-art methods. In particular, their capability to learn discriminative features when trained in a supervised fashion, provides an automated approach for regional assessment of radiographic features. CNN models are being explored extensively for their potential to identify features, sometimes unrecognizable to the naked eye, that correlate to disease outcome. For instance, Cheplygina *et al*. [22] reported a machine learning model using transfer learning for multicenter classification of COPD; Athimopoulos *et al*. [23] and Shin *et al*. [24] proposed CNN models to classify lung patterns for interstitial lung diseases (ILDs) and emphysema, respectively.

To improve the automated detection and quantification of AT that better matches thoracic radiologist visual assessment of AT on expiratory CT images, and to reduce the discordance that can occur between these methods, we trained a CNN model capable of delineating regional AT on expiratory CT images to identify and quantify AT. We further validated this approach through comparison to radiologist CT image assessment and correlation to PFT and clinical scores.

## MATERIALS AND METHODS

### Ethics Statement

The prospective multi-center study was carried out in 36 subjects enrolled in the Novartis CF Natural History Study [25] from 2007–2011 and was approved by the Institutional Review Boards of Site 1 (IRB #6218) and Site 2 (IRB #07-00207). Informed written consent for examination and further data processing was obtained from all patients or legal guardians prior to inclusion.

### Study Participants

CF subjects were school-age children accrued as part of the Novartis/CF Foundation Therapeutics multicenter prospective 2-year natural history study [26]. CT and clinical data were acquired from two different sites, referred to as Site 1 (N=24) and Site 2 (N=12) (**Table 1**), with baseline and follow-up examinations at 3, 12, and 24 months. All chest CT scans were obtained with the same CT quality assurance (QA) protocol using spirometer controlled acquisition of spiral chest CT scans at both institutions. CF subjects were extensively characterized at baseline based on age, gender, height, weight, body mass index, pulmonary function tests (PFT), and radiologic scores. All chest CT scans were obtained when patients were clinically stable without oral or intravenous antibiotics for a pulmonary exacerbation. All subjects had a confirmed diagnosis of CF by pilocarpine iontophoresis sweat chloride testing and CF gene mutation analysis. The CF data has been used earlier to demonstrate the effects that CT registration has on quantifying air-trapping [27], and airway measurements [28] as well as quantifying lobar segmentation [25, 29] to show that clinical measures such as mucus plugging and bronchiectasis are important in assessing childhood CF. Here, we use this data and propose a CNN model to quantify AT.

**Table 1:** Subject Baseline Characteristics at Each Site

|  | Site 1 | Site 2 |
| --- | --- | --- |
| **Number of Cases (N)** | 24 | 12 |
| **Age (yrs)** | 12 (2.5) | 12 (3.2) |
| **Gender (m/f)** | 11/13 | 5/7 |
| **Height (cm)** | 146 (13.2) | 145 (17.3) |
| **Weight (kg)** | 41.2 (10.5) | 40.0 (13.3) |
| **BMI (kg/m<sup>2</sup>)** | 18.9 (2.1) | 18.3 (2.5) |
| **FEV1 (% predicted)** | 99 (10.5) | 94 (10.1) |
| **FVC (% Predicted)** | 102 (12.3) | 101 (7.1) |
| **FEF25-75 (% predicted)** | 94 (19.9) | 82 (25.3) |

Note: Data presented as the mean (standard deviation). BMI is the body mass index. FEV1 is the forced expiratory volume at 1 sec. FVC is the forced vital capacity. FEF25-75 is the forced expiratory flow at midexpiratory phase.

### Pulmonary Function Tests and Clinical Scores

PFTs were obtained as per clinical care guidelines for each respective institution. Forced vital capacity (FVC), forced expiratory volume at one second (FEV1), and forced expiratory flow at 25–75% (FEF25-75) were expressed as percent predicted based on Global Lung Initiative normal prediction equations (**Table 1**).

### CT Acquisition Technique

Volumetric helical CT scans of the chest were obtained using spirometer controlled multidetector CT scanners (Siemens Sensation 64, 32 detectors; Siemens Medical Solutions, Malvern, PA) at Site 1 and (GE VCT scanner, 64 detectors; GE Healthcare, Waukesha, WI) at Site 2 [30]. Inspiratory volumetric chest CT scans were obtained at ≥ 95% vital capacity (VC), while expiratory spiral volumetric CT scans were obtained near residual volume [RV] (c. 5% VC) using the spirometer-controlled CT acquisition.

For all subjects, a low dose spiral CT scanning protocol was utilized with 100 kVp and 30 – 50 mAs at Site 1 and 100 kVp and 20 – 40 mAs at Site 2 with slice thickness of 0.6 – 0.625 mm with 50% overlap in lung and soft-tissue kernels (standard). QA protocol was implemented on-site for adequate inflation and deflation levels, absence of significant motion artifacts, and inclusion of all parts of the chest, by a thoracic radiologist. Technical CT QA was done using standard CT phantoms for both the Siemens and GE CT scanners prior to chest CT scanning for baseline, 3, 12 and 24-month testing. In addition, differences in mAs between the 2 sites were standardized prior to the Novartis/CF Foundation Therapeutics 2-year study utilizing a CT airway and parenchymal phantom [31]. The calculated total effective dose for the 4 serial CT scans from Site 1 and Site 2 (baseline, 3 months, 1 year, and 2 years) was 5.4 – 5.6 mSv. This corresponded to an estimated risk of developing cancer of approximately 0.056% [32, 33].

### Attenuation Threshold-Based Quantitation of Air Trapping

Quantitative air trapping (QAT) was determined on expiratory CT data using a slight variation of the previously reported algorithm developed by Goris et al. [9], referred to here as the Personalized Threshold Method (PTM). This approach generates subject-specific thresholds for detecting regions of lung parenchyma with mild-to-severe AT (originally defined QAT_A1_ in [9]). A 3 × 3 × 3 median filter was applied to inspiratory and expiratory CT scans immediately prior to AT classification. The lungs were then automatically segmented using an in-house software developed using MATLAB (MathWorks, Natick, MA), and voxels with HU values > 0 were excluded. The lungs segmentations were visually inspected to make sure there are no errors. The 50^th^ and 90^th^ percentile for the inspiratory CT scan (Y and X, respectively) and difference in the 90th percentile values in the inspiratory and expiratory CT scans (D) were determined. These HU values were used to calculate a subject-specific threshold (T) for AT using the following expression: T = X − (1 − *D*/343)* (X − Y)/3 [9]. An AT map was generated by classifying all expiratory CT voxels with HU values < T as 1, and the remaining voxels as 0. QAT_PTM_ was calculated from the AT map by summing the binary value of all voxels and normalized to the total number of voxels within the segmentation map (i.e. whole-lung). For completeness, the accepted static threshold (T) of −856 HU was applied to all expiratory CT scans to also quantify AT (QAT_-856_) [13]. For reference, standard air and water attenuation values are −1000 and 0 HU, respectively.

### CNN Algorithm Development

We developed a feature-based method using a densely connected CNN (DN) for detection and quantification of AT. We adopted the CNN model proposed by Huang *et al*. [34] as a basis to construct our DN architecture. The motivation behind this architecture is that the contraction and expansion paths of the architecture captures the context around the objects to provide a better representation of areas of AT on CT images. In our implementation each dense block layer is composed of batch normalization (BN), followed by a rectified linear unit (ReLU) activation, and a 3 × 3 convolution. Each downsampling path consists of BN, followed by ReLU activation, a 1× 1 convolution, and a non-overlapping 2 × 2 max pooling. Also, each upsampling path is composed of a 3 × 3 transposed convolution with a stride of 2 to compensate for the pooling operation.

Our deep learning architecture consists of two downsampling and two upsampling paths, with four dense block layers between each downsampling and upsampling path. After the last upsampling path we perform an 1 × 1 convolution, followed by a softmax operation in order to obtain the final output label for each pixel within the image. A schematic representation of our proposed DN is shown in **Fig. 1**.

**Figure 1:**
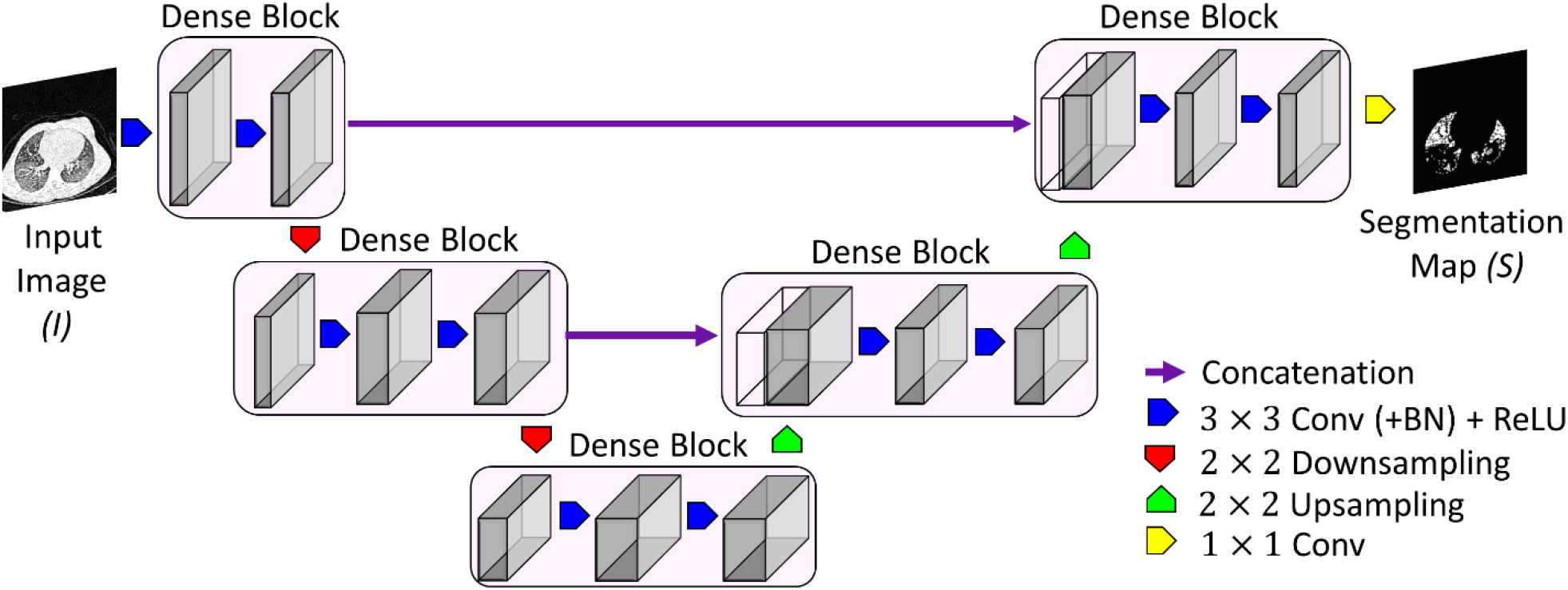
Schematic overview of the proposed dense convolutional network (DN) architecture.

#### Training

The DN was trained on a desktop workstation running a 64-bit Windows operating system (Windows 10) with an Intel Xeon W-2123 CPU at 3.6GHz with 64GB DDR4 RAM and an NVIDIA GeForce RTX 2080 graphic card with 2944 CUDA cores (Nvidia driver 411.63) and 12GB GDDR6 RAM. Our proposed DN architecture was trained to minimize the Dice loss [35, 36]. The x, y, and z-dimensions of each image in our dataset was x = 512, y = 512, and z ∼ 850. We used a randomly selected subset of 22,784 2D slices from 32 3D images (N = 8; with four different time points) from the Site 1 cohort consisting of a total of 69,137 2D slices from 96 3D images (N = 24; with four different time points) for training the DN model. A separate set of 19,592 2D slices from another 32 3D images (N = 8; with four different time points) from the Site 1 cohort were held out for validation and parameter tuning. AT segmentation maps generated using the PTM was used for training the DN. We used a nested 2-fold cross-validation strategy for training the DN architecture, where the outer loop was run eight times and the data was split into two equal random pools internally. The network was trained on one pool and tested on the other.

The proposed DN architecture was implemented in PyTorch [37] and run under the Python environment (version 3.7; Python Software Foundation, Wilmington, Del; https://www.python.org/). We used the stochastic gradient descent algorithm, called Momentum [38], to efficiently optimize the weights of the DN. The weights were normalized using a normal random initialization and updated in a mini-batch scheme of 16 candidates, with a growth rate of 8 per iteration. The biases were initialized to zero, the momentum term was set to *γ* = 0.9, and the learning rate was set to α = 0.001.

### Radiographic Assessment of Air Trapping on CT

Four subjects from Site 1 and two from Site 2 were randomly selected for the visual assessment of AT from expiratory CT images. Including all CT examinations, there were a total of 24 expiratory CT examinations, which were examined by three trained thoracic radiologists. Expiratory CT examinations were loaded onto a laptop with an in-house image viewer developed using MATLAB (MathWorks, Natick, MA) capable of manually applying a threshold to the CT data that generates an overlay indicating regions of lung parenchyma less than the threshold on the CT scan. The threshold was adjusted manually by the radiologist. Once the radiologist deemed the threshold sufficient to highlight AT on a specific expiratory CT scan, the threshold was recorded and used to calculate the QAT for the CT examination. A single preset threshold was not used for all of the four subjects. The radiologists were allowed to vary the window level combination when reviewing the images as each of them had their comfort window level for each image. No additional instructions were given to complete this task. This process was performed separately by all three radiologists and repeated for all 24 expiratory CT scans. QAT values were then averaged over all radiologists for a given expiratory CT scan. Individual averaged results for each of the six subjects are presented in **Table 2**.

**Table 2:** Computed QAT Values from Multiple Radiologist Verified Thresholds and the PTM

| Case ID Number | Time Point | % Quantitative Air Trapping (QAT) Values |  |  | QAT<sub>PTM</sub> |
| --- | --- | --- | --- | --- | --- |
|  |  | Radiologist 1 | Radiologist 2 | Radiologist 3 |  |
| 1 | Baseline | 7.4 | 6.8 | 5.9 | 4.1 |
|  | 3 months | 9.9 | 8.7 | 9.1 | 8.1 |
|  | 1 year | 12.9 | 11.7 | 11.7 | 8.2 |
|  | 2 year | 15.9 | 14.9 | 10.2 | 9.3 |
| 2 | Baseline | 12.8 | 12.1 | 9.3 | 6.5 |
|  | 3 months | 11.2 | 9.9 | 12.3 | 9.3 |
|  | 1 year | 14.9 | 14.6 | 8.2 | 12.4 |
|  | 2 year | 17.1 | 18.7 | 11.2 | 9.6 |
| 3 | Baseline | 6.9 | 8.0 | 5.1 | 4.8 |
|  | 3 months | 9.5 | 8.8 | 5.9 | 10.4 |
|  | 1 year | 12.8 | 12.1 | 7.6 | 10.2 |
|  | 2 year | 13.4 | 12.3 | 10.2 | 9.8 |
| 4 | Baseline | 8.2 | 7.1 | 9.5 | 7.4 |
|  | 3 months | 11.7 | 12.2 | 9.9 | 11.2 |
|  | 1 year | 13.1 | 12.7 | 9.4 | 12.8 |
|  | 2 year | 14.1 | 14.1 | 14.1 | 10.2 |
| 5 | Baseline | 7.5 | 6.6 | 4.5 | 6.1 |
|  | 3 months | 10.1 | 11.4 | 6.3 | 9.6 |
|  | 1 year | 12.4 | 12.4 | 8.1 | 7.2 |
|  | 2 year | 14.9 | 15.6 | 9.7 | 8.4 |
| 6 | Baseline | 8.4 | 7.3 | 6.8 | 9.4 |
|  | 3 months | 11.5 | 12.6 | 8.5 | 11.4 |
|  | 1 year | 14.7 | 13.9 | 18.9 | 10.1 |
|  | 2 year | 15.6 | 16.7 | 12.1 | 9.8 |

### Simulation of Deflation-Levels on Expiration CT Scan

Adequate respiratory maneuvers are of importance in CT imaging of patients when assessing air trapping over time [39]. Also, there is no consensus on how the expiratory CT data is acquired for measurement of AT [14]. To address these concerns on AT quantification using expiratory CT scans, we studied the effects of lung deflation on the quantification of AT determined using our CNN and PTM. Inadequate lung deflation on the expiratory CT acquisition was simulated using paired CT data from 4 subjects, two with AT < 10% total lung volume and two with AT > 25% total lung volume, as defined by QAT_PTM_. The inspiratory CT acquisition data was registered to the expiratory CT data for each subject at a single time point (1 registration per case). All registrations were performed using Elastix (version 4.8), an open-source deformable image registration library [40, 41]. We used the B-spline non-rigid transformation to register the inspiratory CT acquisition data to the expiratory CT data. This algorithm iteratively optimizes the solution using mutual information with a bending energy penalty as the objective function. Mass was preserved by adjusting the HU values for volume changes by multiplying each voxel by the local determinant of the Jacobian matrix of the warping transform. Simulated expiratory CT acquisitions were generated assuming a linear trajectory between the expiration and inspiration lungs. The determinant of the Jacobian matrix was linearly altered to reflect different deflation levels using the following expressions:

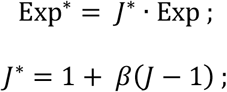

where *J* and *J*^*\**^ are the original and simulated determinant of the Jacobian matrix, Exp and Exp^*\**^ are the original and simulated expiratory CT scan, and *β* is the fraction deflated, such that *J*^*\**^ = 1at no deflation (*β* = 0) and *J*^*\**^ = *J* at full deflation (*β* = 1). QAT measurements using PTM and DN were determined on the original (*β* = 1) and simulated expiration CT data (*β* = 0.9, 0.8, 0.7, 0.6, and 0.5).

### Statistical Analysis

All data values are presented as the mean ± standard deviation. Site comparisons were determined for patient characteristics, and PFT measure using an unpaired Wilcoxon test. Wilcoxon signed rank test was used to assess differences in the quantitative AT measurements, PFT, and clinical scores between interval examinations. QAT measures using DN were correlated to PFT and clinical scores using linear mixed effects modeling. We used a fixed effect term at the population level (i.e., the entire cohort) and a correlated random effect grouped by each subject within the cohort for the 4 times each subject was measured. Results were considered statistically significant at the 2-sided 5% comparison-wise significance level (p < .05). All statistical computations were performed with a statistical software package (IBM SPSS Statistics, v. 21, Armonk, NY).

## RESULTS

### Study Patient Demographics

Negligible differences in patient characteristics and pulmonary function were observed from the clinically confirmed mild CF patients accrued at the two sites (**Table 1**). Baseline QAT measurement using a hard threshold of −856 HU (QAT_-856_) was 3.53% ± 1.29%. When applying the PTM approach to quantify AT, QAT values were nearly double when compared to using the static threshold to quantify AT (QAT_PTM_; 6.79% ± 1.83%).

### Comparison of QAT Measurements

Illustrated in **Fig. 2**, is a representative axial 2D slice from a female in her 20’s diagnosed with mild CF (FEV1% predicted value of 96%) at the 12-month examination from the Site 1 cohort. No clear mosaic patterns were identified radiographically on the inspiratory CT images (**Fig. 2 A**). In contrast, a mosaic attenuation pattern is present on the expiratory CT images indicating AT (**Fig. 2 B-D**). Applying a hard threshold of −856HU captured 3.8% of the total lung volume as AT, which increased to 7.8% using the PTM approach (threshold adjusted to −787 HU). QAT values determined using the DN model were in agreement with the radiologist visual assessment, with QAT values around 10% (**Fig. 2 D-E**). The mean radiologist threshold determined by visual review to be sufficient to identify most of the AT for this specific CT scan was −726 HU.

**Figure 2:**
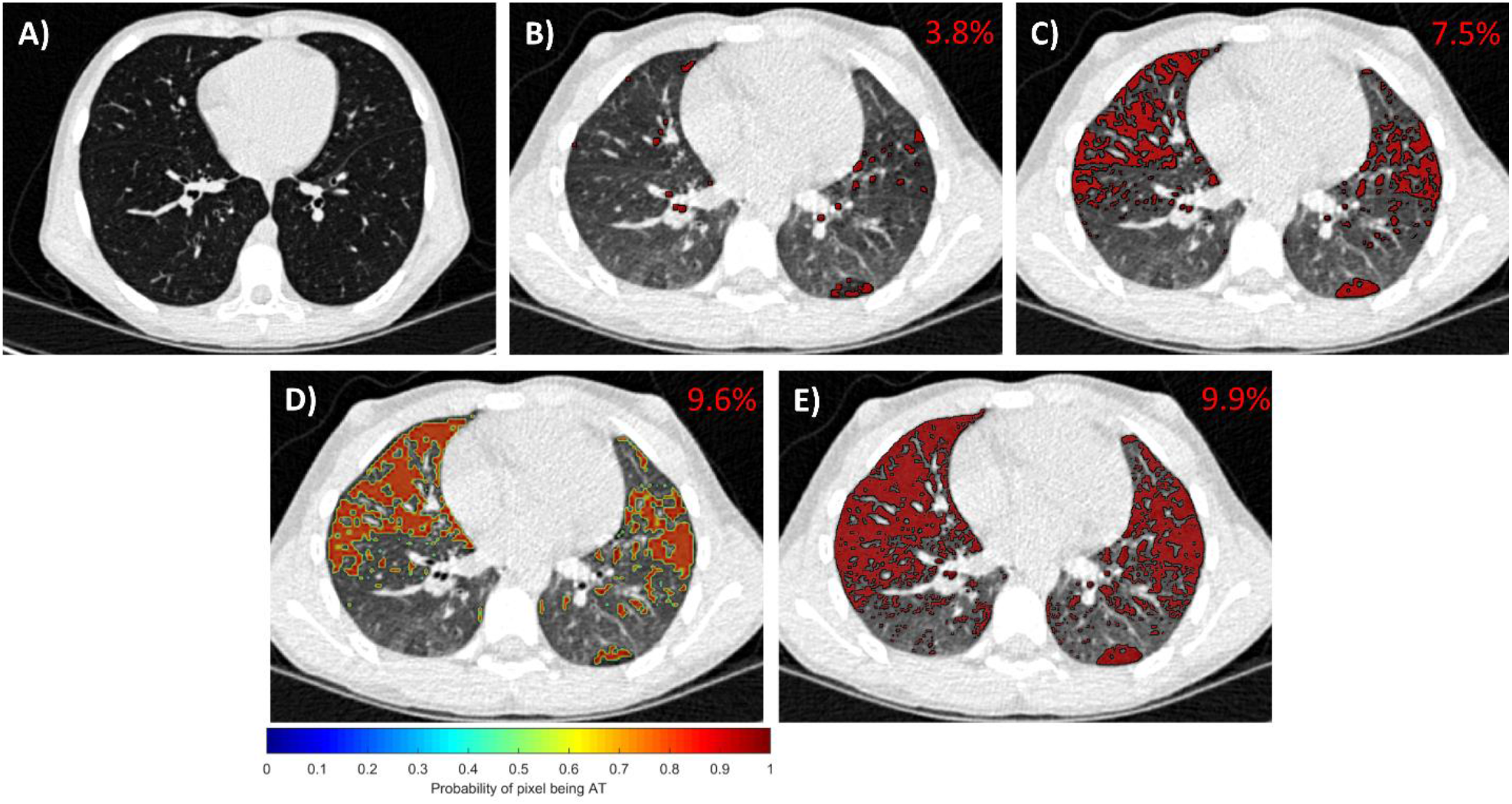
Presented from a single case from the Site 1 examined at 12 months are representative images from A) the inspiratory CT scan and AT maps from B) a static threshold of −856HU, C) the personalized threshold method (PTM), D) the DN method (presented as probability map), and E) radiographic assessment by a trained radiologist. The QAT for the whole 3D dataset is provided on the top right corner of each 2D image. The images are windowed between [-950, 150] for display purposes so that regions of AT are visible.

The DN model was found to detect AT that increased in a time dependent manner. At baseline, good agreement was observed for the QAT_DN_ to QAT_PTM_ with a difference in QAT value of about only 1.2% (**Fig. 3**). Evaluating the QAT values over time, the poor agreement between QAT_DN_ values to QAT_PTM_ was attributed to the ability of the DN model to detect increasing amounts of AT over the two-year period (**Fig. 3**). Follow-up QAT_PTM_ were found to be significantly higher at about 1.5 times the baseline values. Nevertheless, these values plateaued with no significant difference between interval QAT_PTM_ measurements post-baseline examination with a *p-*value of 0.25 and 0.73 between the intervals of 3 to 12 months and 12 to 24 months, respectively. The QAT_DN_ significantly increased from baseline to year two by up to 12.7% ± 1.1%. The QAT_DN_ measurements post-baseline examination were also significantly different with a *p*-value of 0.004 and 0.003 between the intervals of 3 to 12 months and 12 to 24 months, respectively. Radiologist visual assessment and threshold setting showed similar trends in QAT to values obtained using the DN model, although not with the same rate of incline (**Fig. 3**). Mean QAT_-856_ values at baseline, 3months, 1 year, and 2-year were found to be 4.3% ± 2.6%, 4.9% ± 3.8%, 5.3% ± 2.9%, and 4.6% ± 1.9%, respectively. None of the follow up QAT_-856_ values were found to be significant to the baseline value.

**Figure 3:**
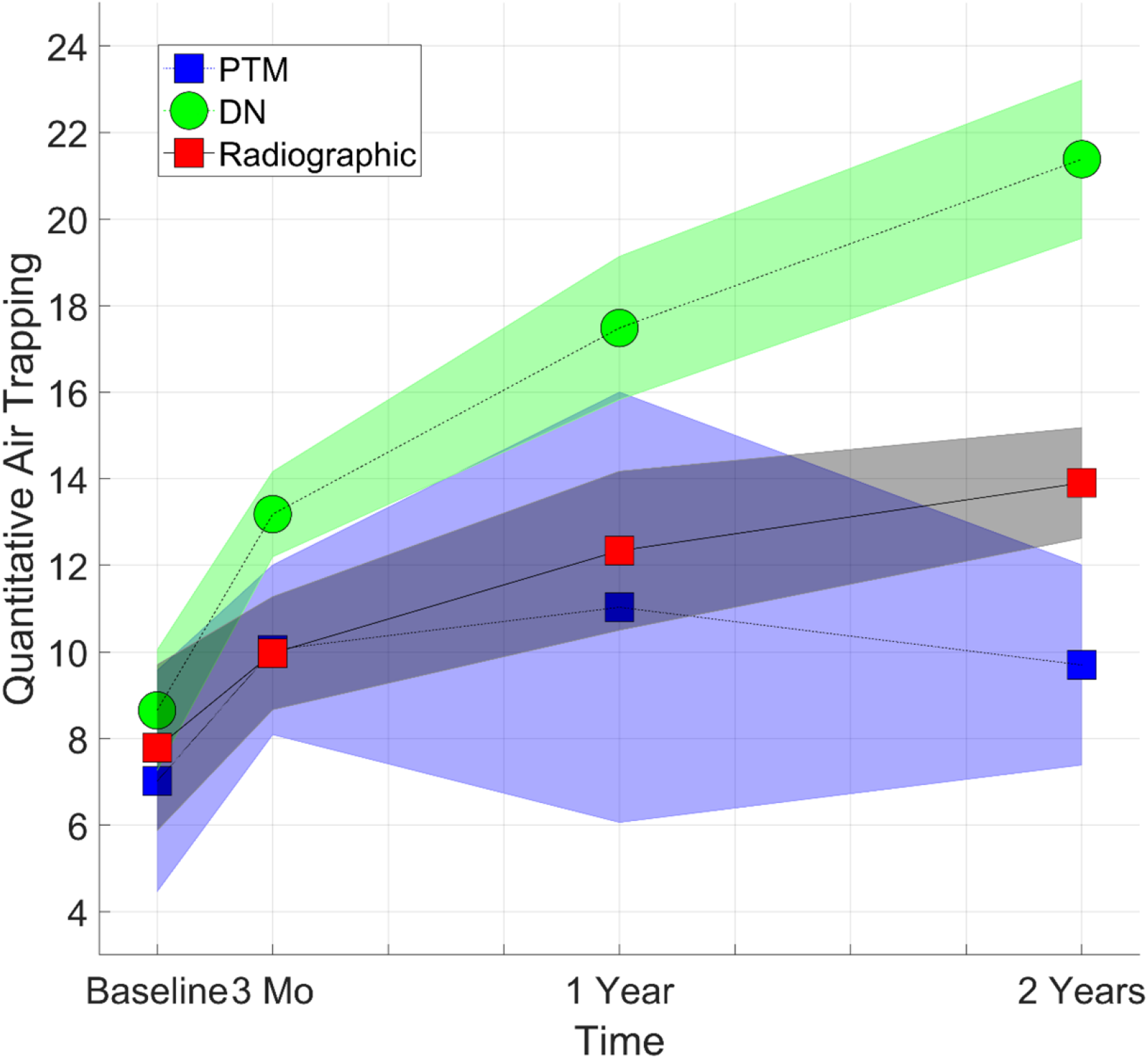
Presented are Quantitative air trapping (QAT) results of the various methods from Site 1 cohort at the different examination times.

### Multi-Site Comparison

To validate the robustness and generalizability of the CNN approach, QAT values generated from the fully-trained DN model at the Site 1 cohort were evaluated using CT examinations from the Site 2 cohort. A representative axial CT slice from a young boy in the age group 10-20 with CF (FEV1% predicted value of 92%) at the 3-month examination showed similar QAT values for PTM, DN, and visual threshold assessment (**Fig. 4A**). Peripheral AT on the dependent posterior regions of the lungs was undetected by PTM (threshold value of −802 HU), and was detected by the DN model and also confirmed by radiographic assessment (RA) (threshold value of −743 HU). As observed from the Site 1 cohort, QAT_PTM_ values plateaued after the 3-month examination, with both QAT_DN_ and QAT_RA_ significantly increasing up to 6.8% ± 0.2% and 4.9% ± 0.58%, respectively, by year two (**Fig. 4B**). The interval QAT_DN_ measurements post-baseline were significantly different with *p*-values of 0.03 and 0.04 between 3 to 12 months and 12 to 24 months, respectively. Mean QAT_-856_ values at baseline, 3 months, 1 year, and 2-year were found to be 4.8% ± 1.9%, 5.8% ± 2.5%, 6.1% ± 3.7%, and 5.7% ± 2.8%, respectively. None of the follow up QAT_-856_ values were found to be significant to the baseline value.

**Figure 4:**
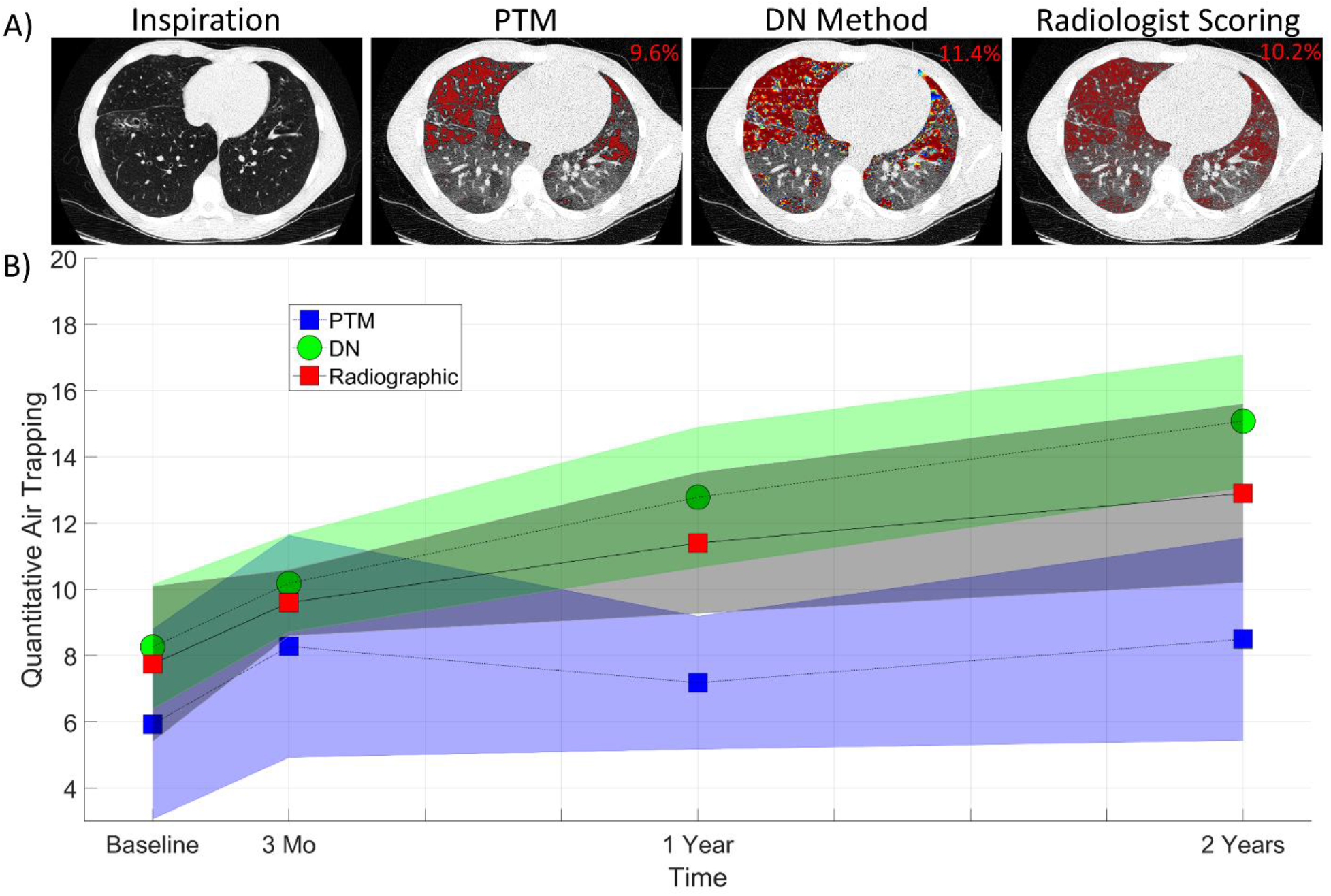
A) Presented from a single case from the Site 2 cohort examined at 3 months are representative images from A) the inspiratory CT scan and the AT maps from the personalized threshold method (PTM), the DN method, and radiographic assessment by a trained radiologist. The QAT for the whole 3D dataset is provided on the top right corner of each 2D image. The images are windowed between [-950, 150] for display purposes so that regions of AT are visible. B) Quantitative air trapping (QAT) at different times by the various methods on the Site 2 cohort.

### Radiologist Visual Assessment

Radiologist visual assessment and threshold setting showed similar trends in QAT values to that observed by our DN model, although not with the same rate of incline. In general, AT by radiographic assessment was less than as quantified by our CNN models. We did observe lower AT values from Radiologist 3 from what was reported from Radiologists 1 and 2 (**Table 2**).

### PFT and Clinical Scoring

Pulmonary function measurements and clinical scores were assessed over the entire study population (N=36), to determine if trends in these clinically relevant outcomes were similar to those observed in the QAT values measured by the DN model. Although all mean PFT measures steadily decreased, follow-up values were not significantly decreased from baseline. Nevertheless, these trends suggest pulmonary dysfunction, although variable from case-to-case, throughout the duration of the study (**Table 3**). In contrast, radiologic scores for AT, bronchiectasis, and mucus plugging, all demonstrated a significant increase at year two. The most pronounced increase was observed for the mucus plugging score, which increased by nearly 140% and was found to significantly increase at all follow-up examination time points (**Table 3**).

**Table 3:** Mean and (Standard Deviation) of Pulmonary Function Test and Clinical CT Scores

| Ventilation | Parameter | Baseline | 3 Months | 1 Year | 2 Year |
| --- | --- | --- | --- | --- | --- |
| PFT | FEV1 % Predicted | 93.54 (7.42) | 93.01 (9.01) | 91.69 (9.95) | 87.85 (10.03) $p = 0.06$ |
| | FVC % Predicted | 101.44 (10.56) | 99.89 (11.70) | 99.67 (11.12) | 95.39 (14.69) $p = 0.09$ |
|  | FEF25-75 % predicted | 90.30 (21.93) | 87.83 (26.71) | 85.47 (22.67) | 83.61 (20.80) |
| Clinical Scores | Brody ATS | 26.19 (13.52) | 29.81 (16.10) | 30.58 (14.66) | 33.10 (14.36) * |
|  | Brody BS | 5.84 (8.21) | 5.46 (7.37) | 5.93 (6.67) | 7.29 (7.20)* |
|  | Brody MPS | 2.90 (5.04) | 3.63 (5.36)* | 5.87 (7.28)* | 7.03 (8.06)* |

Note: Pairwise differences in the follow-up PFT measurements and clinical scores from the baseline were tested using the non-parametric Wilcoxon rank sum test. Significance at the 0.05 $p$ -value are indicated as \*. $P$ -values are stated for $p < 0.1$ in the table. Brody ATS = Brody air trapping score, Brody BS = Brody bronchiectasis score, and Brody MPS = Brody mucus plugging score. We used all subjects (N = 36) from both Site 1 and Site 2 in this study.

The PFT and clinical scores were correlated with the QAT values of the DN model using linear mixed effects modeling (**Table 4**). We observed a good correlation between the FVC % predicted and the percentage of AT computed using our DN model. We also observed that the Brody BS and Brody MPS clinical scores very strongly correlated with the QAT_DN_ values. In contrast, the QAT_PTM_ and the QAT_-856_ values did not correlate well with the PFT and clinical scores (**Table 4**).

**Table 4:**
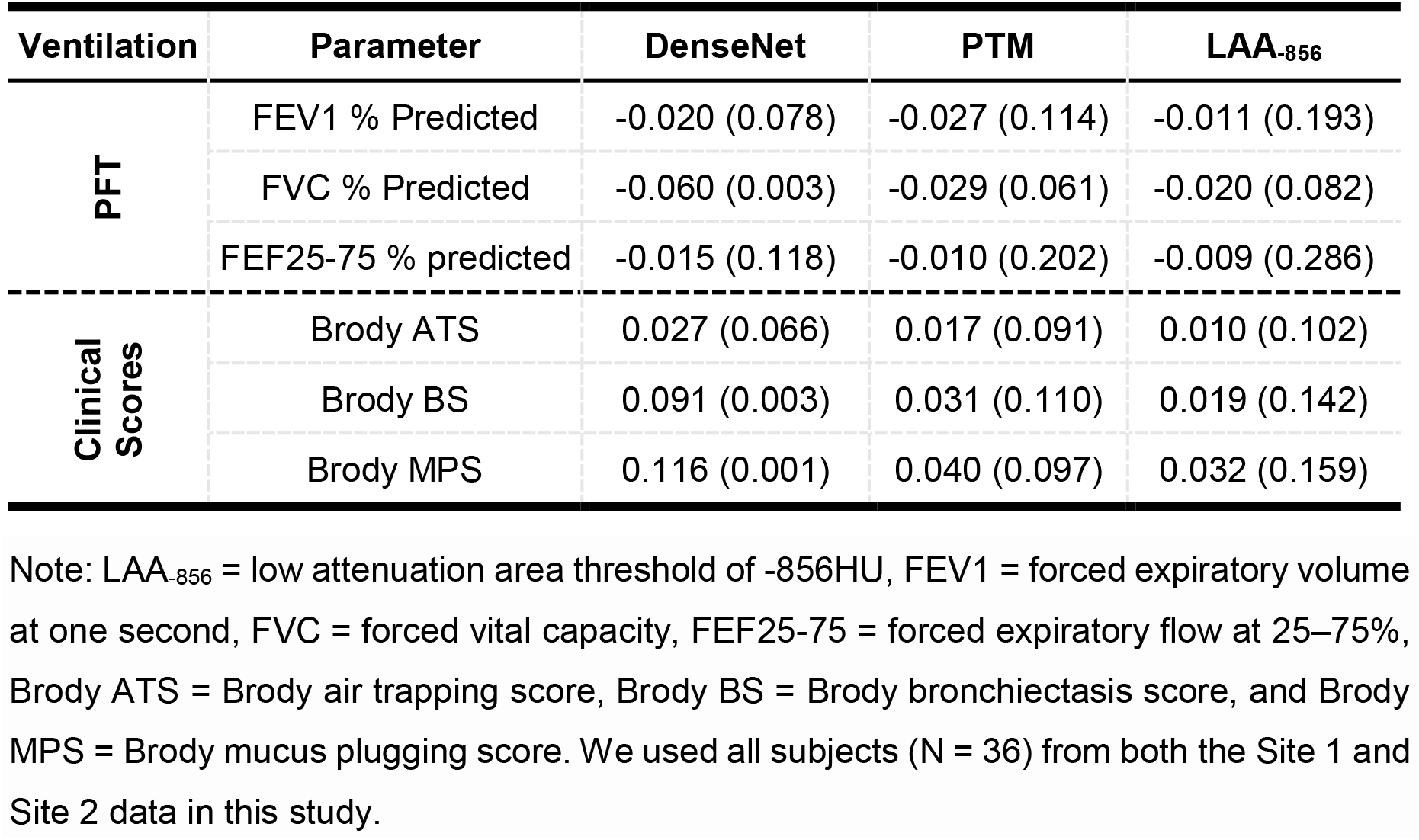
Regression Coefficient and (*p*-values) of Pulmonary Function Tests and Clinical CT Scores using Linear Mixed-Effects Model

| Ventilation | Parameter | DenseNet | PTM | LAA<sub>-856</sub> |
| --- | --- | --- | --- | --- |
| PFT | FEV1 % Predicted | -0.020 (0.078) | -0.027 (0.114) | -0.011 (0.193) |
|  | FVC % Predicted | -0.060 (0.003) | -0.029 (0.061) | -0.020 (0.082) |
|  | FEF25-75 % predicted | -0.015 (0.118) | -0.010 (0.202) | -0.009 (0.286) |
| Clinical Scores | Brody ATS | 0.027 (0.066) | 0.017 (0.091) | 0.010 (0.102) |
|  | Brody BS | 0.091 (0.003) | 0.031 (0.110) | 0.019 (0.142) |
|  | Brody MPS | 0.116 (0.001) | 0.040 (0.097) | 0.032 (0.159) |

Note: LAA<sub>-856</sub> = low attenuation area threshold of -856HU, FEV1 = forced expiratory volume at one second, FVC = forced vital capacity, FEF25-75 = forced expiratory flow at 25–75%, Brody ATS = Brody air trapping score, Brody BS = Brody bronchiectasis score, and Brody MPS = Brody mucus plugging score. We used all subjects (N = 36) from both the Site 1 and Site 2 data in this study.

### Effect of Deflation-Level

Quantitative AT techniques are highly susceptible to the deflation-level executed during the expiratory CT scan. To test this effect on the ability of the DN model to detect AT, HU values from four randomly selected expiratory CT acquisitions from the 144 scans (36 cases * 4 time points) were adjusted to simulate inadequate deflation-levels. Illustrated in **Fig. 5A** is a representative axial CT slice acquired at residual volume (RV) and their corresponding simulated CT image at deflation-levels between 90% to 50% of RV. For demonstration, the CT scan was from a young girl in the age group 5-10 with CF (FEV1% predicted value of 103%) at 3-month time period. As expected, mean HU values dropped from −288HU at RV to −331HU at 50% of RV (**Fig. 5A**). The QAT_PTM_ and QAT_DN_ values from the three month expiratory CT acquisition were 6.3% and 12.1%, respectively. QAT values from QAT_PTM_ and QAT_DN_ methods increased to 63.2% and 44.9% at a deflation-level of 50% of RV, respectively. From the four selected cases, the differences in lung volume from TLC to RV on average were 1.95L ± 0.48L (**Fig. 5B**). As expected, QAT_PTM_ values, the intensity-based technique, were highly sensitive to slight deviation from RV. At a simulated deflation-level of 80% of RV, changes in lung parenchymal density resulted in 15% more AT as measured by PTM, increasing linearly with deflation-level (**Fig. 5C**). Although the DN model, a feature-based technique, measured increasing levels of AT with decreasing deflation-level, the deviation rate was slower than what was observed for the PTM method. At 80% RV, QAT_DN_ measured only 5% ± 2.3% more AT (**Fig. 5C**). For completeness we computed the increase in the QAT_-856_ values from the expiratory CT acquisition. The increase in the QAT values were 7.1% ± 1.9%, 21.4% ± 6.7%, 43.7% ± 5.6%, 60.8% ± 5.1%, 74.5% ± 4.8% at deflation levels between 90% and 50% of RV. The QAT_-856_ values showed the highest sensitivity with respect to increasing levels of AT with decreasing deflation-levels.

**Figure 5:**
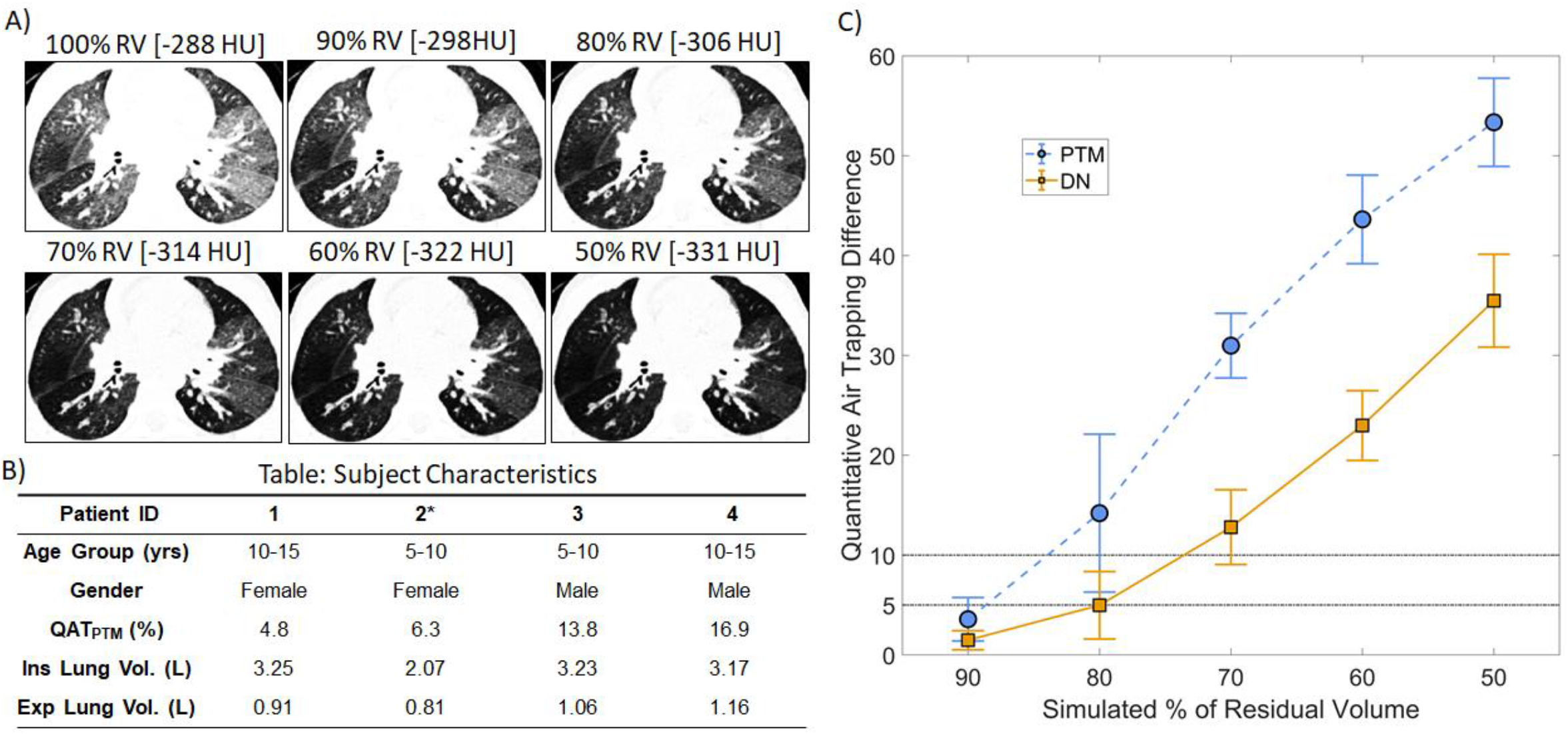
Presented are the results of the effect of deflation level on the expiratory CT scans in quantifying AT. A) A representative image of the expiratory CT scan from a single case (from Patient 2*) from the Site 1 cohort examined at baseline and its corresponding simulated % of residual volume (RV) 2D images. The images are windowed between [-1000, −50] for display purposes so that regions of AT are visible. [·] represents the mean intensity within the whole lung region in HU. B) Patient characteristics table for the four cases considered in this study. C) Plot of average quantitative air trapping (QAT) difference VS the simulated % of RV for the PTM, and DN method.

## DISCUSSION

We set out to demonstrate the utility of our DN model to more accurately quantify the extent of AT on chest CT image acquisitions in a cohort of pediatric CF patients. These cases were accrued as part of a prospective natural history study from two CF centers and comprised of four chest CT examinations over a two-year period. Cystic Fibrosis is AT specific and has unique features that relate well to modeling a CNN. The personalized threshold method (PTM), that we used for training our CNN model was built on the CF patient data. We wanted to focus on an airway dominant disease and thus chose to evaluate patients with CF. Unlike attenuation threshold-based techniques, we observed a significant increase in quantitative AT over the duration of the trial with the DN model. These findings were in concordance with radiologist visual assessment and with mucus plugging scores. With the ability of our DN model to detect unique features, our model was less prone to errors associated with insufficient exhalation during expiratory CT acquisitions than a threshold-based technique, and therefore may not require a correction factor for differing deflation maneuvers over the course of four different testing periods as was needed by Robinson and colleagues [26].

To fully appreciate the ability of our DN model to quantify AT on expiratory CT acquisitions, we elected to use a cohort of pediatric patients. Although observational trials, such as COPDGene [13] and SPIROMICS [42] are at the forefront of advancing quantitative CT algorithms, techniques developed from these older populations fall short when applied to pediatric patients due to the physiological differences in the lungs of these age groups. In general, younger patient lungs tend to have higher attenuation values than older patient lungs, as the residual volume of lungs increases with age [43, 44]. Also, techniques such as classifying lung parenchymal regions based on a static threshold, such as −856 HU, do not always capture the extent of AT that is visible on CT images as demonstrated in **Fig. 1 B**. In addition, we observed interobserver variability in the radiographic assessment of AT between the three Radiologists of this study (**Table 2**). With the ability to identify higher order features, our DN model provides a unique opportunity to overcome these deficiencies to accurately and objectively quantify AT.

It is important to note the limitations of this study. First, our DN model was trained using a patient specific quantitative algorithm (PTM) for segmenting AT on the expiration CT acquisition, rather than the conventional approach that requires a trained radiologist to contour the air trapped regions, a time consuming and arduous process. In an effort to accelerate this process, AT segmentation maps from the PTM were explored as an alternative. This proved advantageous, as we were able to fully automate the training set and using sub-optimal segmentation maps (i.e., segmentation maps with noisy labels) proved to possess sufficient information to train the DN model. A second limitation was the small number of cases in the cohorts, which may have introduced bias in the DN model. To address this concern, we trained and tested a separate CNN model called the scattering convolutional network (SN) that is based on a scattering transform defined by Bruna and Mallet [45], using the same method (see **S1 Appendix**). Individual cases from the Site 1 cohort were randomly selected such that the training and testing sets varied for the two architectures (i.e. DN and SN). To avoid a bias in the time of examination, all four time points of the CT data were included for each case in the training and testing sets. We found that the QAT values from the SN model were similar to those of the DN model (**S2 Fig.)**. This strategy provides some degree of reassurance that the observed time dependent increase in the QAT measurements from our DN model is accurate.

In conclusion, we developed a deep learning algorithm that can accurately quantify the extent of AT on chest CT images. We tested our proposed CNN model on a cohort of pediatric CF patients that underwent CT examination four times over a two-year period. Our study found a significant increase in the QAT in these patients over the duration of the trial. To the best of our knowledge, this study is the first to propose a deep learning algorithm to quantify AT in chest CT images. Quantitative AT measured using this method can be used as an imaging biomarker for assessment of disease severity and can aid in the clinical management of patients with diseases such as CF, Asthma, and COPD. Future work will further evaluate the generalizability of this model to additional data sets such as COPDGene and SPIROMICS.

## Data Availability

All relevant data are within the paper and its Supporting Information files.

## Supplementary Material

### SUPPLEMENTARY METHOD 1

#### Scattering Convolutional Network (SN)

To confirm that the results observed using our proposed DN method did not introduce any bias, we trained and tested a separate CNN model referred to as the scattering convolutional network (SN). The SN model was based on a scattering transform defined by Bruna and Mallet in [1]. The proposed SN consists of a total of four scales (***J*** = 4) where at each scale we employ a Gabor wavelet (***W***) with five different orientation filters (*K* = 5) separated by π*k/K* angles, where *k* = {*1*, ⋯, *K*}. The scattering coefficients at each scale were obtained by a wavelet transformation, followed by a non-linear pointwise complex modulus applied to the wavelet coefficients, a local averaging operator ***A***_*J*_, and downsampling of scale 2^*J*^. Once all scattering coefficients were obtained, they were concatenated together and used as features for classification. We trained the model using a generative principal component analysis (PCA) classifier using the features from SN to identify the areas of AT on the CT images. A schematic representation of the proposed SN is shown in **S1 Figure**.

**S1 Figure:**
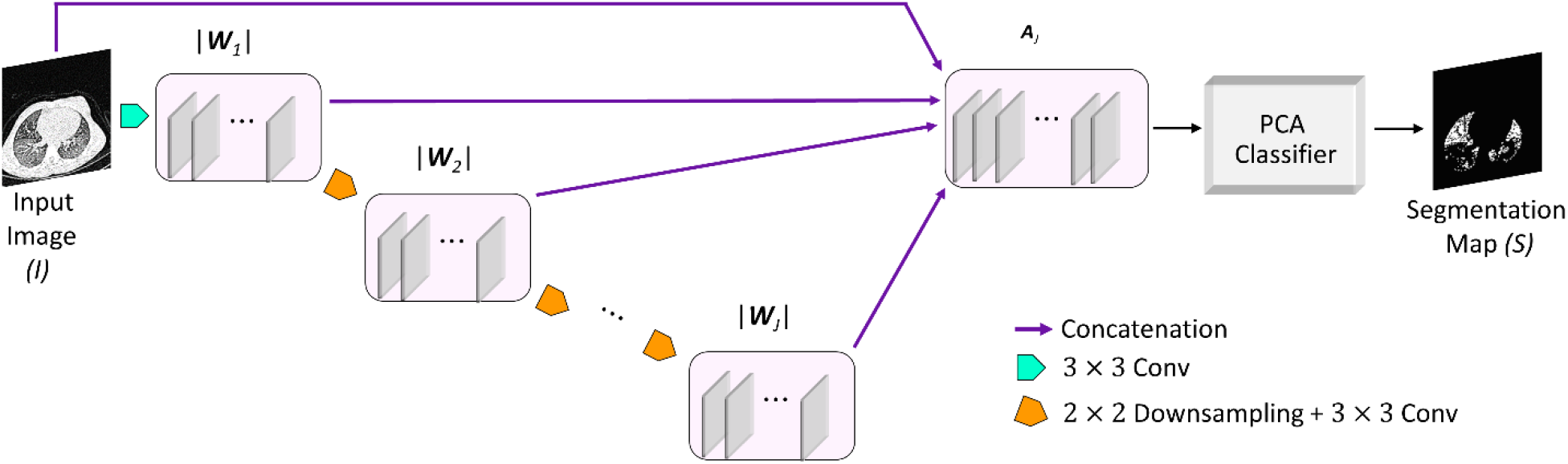
Schematic Overview of SN. Schematic overview of the scattering convolutional network (SN) architecture, where the |***W***_*J*_|’s represent the modulus wavelet transform at each scale *J*, and ***A***_*J*_ concatenates the averaged signals (the detailed coefficients) of the wavelet transform at all scales. We used a total of *J* = 4 scales in our implementation.

##### Training

The SN was trained to minimize the Dice loss, similar to that of DN. Similar to the DN model a nested 2-fold cross-validation strategy was employed for training the SN model. Although, we used the same strategy for training both the SN and the DN model using the data from the Site 1 of 96 images (N = 24; with four different time points), the training and test data were randomly selected for each model separately. The SN architecture was implemented in MATLAB (version 2019a, MathWorks, Natick, Mass). The filters in the SN architecture are fixed Gabor wavelet filters at different orientations. The final layer in the SN architecture was trained using a PCA classifier as described in [1]. The SN was trained on a workstation different from that of the DN method running a 64-bit Windows operating system (Windows 10) with an Intel Xeon W-2123 CPU at 3.6GHz with 64GB DDR4 RAM and an NVIDIA GeForce RTX 2080 graphic card with 2944 CUDA cores (Nvidia driver 411.63) and 12GB GDDR6 RAM.

### SUPPLEMENTARY RESULT 1

#### Comparison of QAT Measurements

The SN model was found to detect air trapping that increased in a time dependent manner similar to the DN model. At baseline, good agreement was observed for the QAT_SN_ to QAT_PTM_ with a difference in QAT values of about only 1% (**S2 Fig**.). Evaluating the QAT values over time, we saw a poor agreement between the SN QAT values to the QAT_PTM_ as the SN model detected increasing amounts of AT over the two-year period (**S2 Fig**.) like that of the DN model in **Fig. 3**). The QAT_SN_ significantly increased from baseline to year two of the trial by up to 11.6% ± 1.7%. The QAT_SN_ measurements post-baseline examination were also significantly different with a *p*-value of 0.03 between the intervals of 3 to 12 months and a *p-*value of 0.002 between the intervals of 12 to 24 months, respectively.

**S2 Figure:**
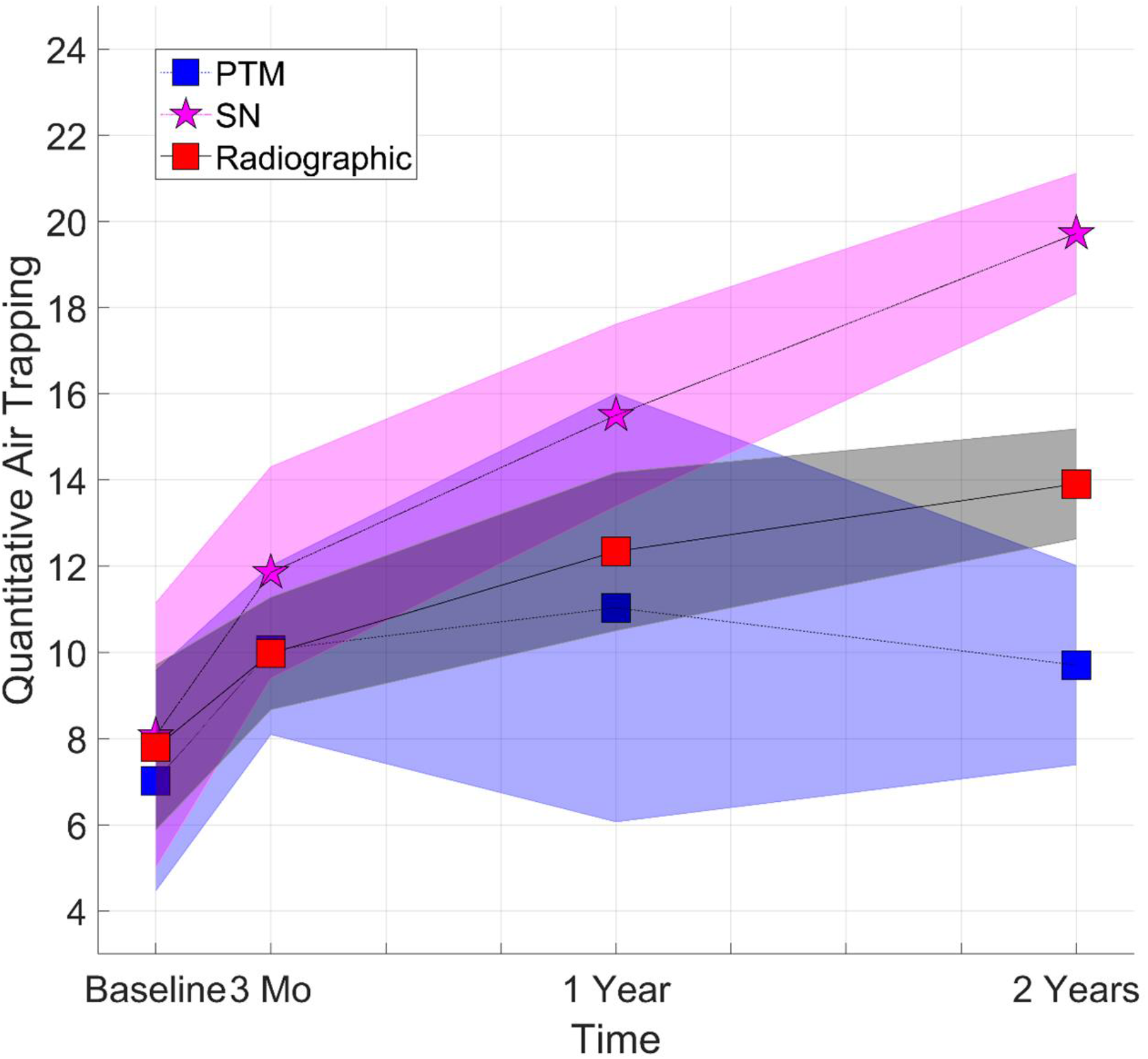
QAT Results for the SN Method. Presented are Quantitative air trapping (QAT) results of the various methods from Site 1 cohort at the different examination times.

